# Menstrual hygiene management in two districts of Malawi

**DOI:** 10.1101/2024.04.12.24305724

**Authors:** Rebekah G.K. Hinton, Laurent-Charles Tremblay-Lévesque, Modesta Kanjaye, Christopher J.A. Macleod, Mads Troldborg, Robert M. Kalin

## Abstract

Menstrual hygiene management (MHM) forms a critical component of ensuring access to adequate and equitable sanitation for all, as outlined in SDG 6.2. Despite its importance, little is known about MHM in Malawi, particularly at a household level. Through a household survey of MHM within 2 districts, we evaluated the type of menstrual absorbents used by people who menstruate. Reusable cloths/rags were the most used menstrual absorbent, used by 79.5% of respondents, whilst disposable absorbents, such as tampons and sanitary pads, were used by 18.6% of respondents. Appropriate MHM also incorporates adequate management of MHM materials, including the washing and drying of reusable menstrual absorbents. We evaluated the cleaning of reusable menstrual absorbents; most respondents (90.1%) reported appropriate washing of menstrual absorbents using soap and water, however only 20.3% reported that menstrual absorbents were dried outside in the sun (as is best practise) with most reporting that reusable menstrual absorbents were dried inside their homes. Our findings highlight the need for improved MHM within Malawi, not only in the access and affordability of appropriate menstrual absorbents but also the promotion of appropriate washing and drying of menstrual absorbents.

## 2 Introduction

Menstrual hygiene management (MHM) has been defined by the WHO and UNICEF Joint Monitoring Programme as “Women and adolescent girls using a clean menstrual management material to absorb or collect menstrual blood, that can be changed in privacy as often as necessary for the duration of a menstrual period, using soap and water for washing the body as required, and having access to safe and convenient facilities to dispose of used menstrual management materials. They understand the basic facts linked to the menstrual cycle and how to manage it with dignity and without discomfort or fear”(UNICEF, 2019; WHO & UNICEF (JMP), 2012). Whilst ensuring appropriate MHM is not outlined as a specific goal within the SDGs, it is a central component of meeting the hygiene and sanitation needs of people who menstruate, thereby a central component of meeting sustainable development goal 6.2 “To achieve access to adequate and equitable sanitation and hygiene for all” (UN General Assembly). Furthermore, the importance of considering the ‘needs of women and girls’ in achieving equitable sanitation and hygiene is particularly emphasised in SDG6.2 (UN General Assembly). Moreover, appropriate MHM also strongly contributes to the achievement of other SDGs, namely, SGD3: “Good health and wellbeing”, SDG4; “Inclusive and equitable quality education and promote lifelong learning opportunities for all”; SDG5: “gender equality and empower all women and girls”; and SDG8: “Promote sustained, inclusive and sustainable economic growth, full and productive employment and decent work for all” (Assembly, n.d.; Ssewanyana & Bitanihirwe, 2019).

Access to, and use of, appropriate menstrual absorbent materials is a critical part of ensuring appropriate MHM (Phillips-Howard et al., 2016). However, millions of people who menstruate in low- and middle-income countries still struggle to access appropriate MHM (Kambala et al., 2020). The most common menstrual absorbents used in resource-poor countries are “old cloths, tissue paper, cotton or wool pieces, or some combination of these items” (Kuhlmann et al., 2017, p 358). Reusable cloths are often made from bunching up and sewing scraps of old clothing, towels, or blankets (McMahon et al., 2011). In Malawi, rags and cloths used as menstrual absorbents are then often looped with string tied around the waist and further held in place by underwear (Pillitteri, 2011). A study of schoolgirls in rural Uganda identified that 87% were using rags as menstrual absorbents (Boosey et al., 2014) whilst similar studies in Nigeria estimated that 31% and 55.7% of schoolgirls used toilet tissue or cloth as a menstrual absorbent (Adinma et al., 2008; Aniebue et al., 2009).

When rags and cloths are unavailable, some people who menstruate must resort to using plant materials such as soft grasses (Boosey et al., 2014) and leaves (Vaughn, 2013) to provide a menstrual absorbent. In the absence of any appropriate menstrual absorbents, many people who menstruate are forced to isolate (Vaughn, 2013), this has particularly significant consequences for adolescents and has been linked to school absenteeism. One study in Uganda reported that over 60% of the girls surveyed reported missing school each month for menstrual-related reasons (Boosey et al., 2014). A similar study in Ethiopia found that girls who did not use menstrual absorbents were over five times more likely to miss school, some dropped out of school following teasing about blood-stained clothes. Furthermore, 58% of girls reported a decline in school performance after the onset of menstruation (Tegegne & Sisay, 2014). This is a significant consideration in reaching SDG4, achieving “inclusive and equitable quality education”. Commercially available, disposable menstrual absorbents are frequently reported to provide higher absorbency (Foster & Montgomery, 2021; Kambala et al., 2020) and are often a preferred form of menstrual absorbent (Hennegan et al., 2016), particularly by younger users (Kambala et al., 2020). However, many people who menstruate in low-resource countries are unable to use this method due to cost (Boosey et al., 2014; Vaughn, 2013) and may resort to using unsafe materials in menstrual hygiene management (Kambala et al., 2020; Vaughn, 2013).

The 2021 JMP MICS report for Malawi found that 97.3% of women reported having access to appropriate materials for menstruation during their last menstruation with 68.5% of women reporting using reusable menstrual absorbents. Reusable absorbents were more commonly used in rural communities and among older women (NSO, 2021). However, to evaluate whether such reusable absorbent use is appropriate, it is also important to consider the method of cleaning and management of reusable absorbents. Poor menstrual hygiene practices, particularly when using reusable absorbent pads, have been associated with urogenital and reproductive tract infections (Das et al., 2015; House et al., 2012.; Torondel et al., 2018). The method of washing and sanitising reusable menstrual absorbents is a particularly important consideration in assessing menstrual hygiene and health (Mahajan, 2019; Narayan et al., 2001; Ssewanyana & Bitanihirwe, 2019). Reusable menstrual absorbents should be washed using clean water and soap, as well as dried in the sun in order to minimise microbial growth (Das et al., 2015; Torondel et al., 2018). Access to appropriate menstrual hygiene management may also have a significant impact on psychological wellbeing (Roxburgh et al., 2020). Negative feelings towards menstruation are commonly reported (Chandra-Mouli & Patel, 2017; Vaughn, 2013, Roxburgh et al., 2020; Enzler et al., 2018) including “embarrassment, shyness, anxiety, shame, and stigmatization” (Kambala et al., 2020). Inadequate access to appropriate menstrual absorbents furthers such negative emotions; leakages and signs of menstruation are reported as a major cause of embarrassment and stigma surrounding menstruation (Kambala et al., 2020; Vaughn, 2013, Roxburgh et al., 2020).

Effective MHM has significant consequences for the health, education, economic potential, and gender equality for people who menstruate (Ssewanyana & Bitanihirwe, 2019). However, despite its importance, MHM information in Malawi is limited with no specific information on MHM reported in Census, MIS, and DHS surveys (NMCP and ICF, 2018, NSO 2018, NSO and Macro International, 2016). Studies where MHM has been investigated in Malawi often focus on MHM in schools (Grant et al., 2013; Mchenga et al., 2020; Pillitteri, 2011.; Shah et al., 2023). Whilst MHM within school settings in an important area, it is also vital to also evaluate MHM in the wider community (Kambala et al., 2020).

This study evaluates the types of menstrual absorbents used by people who menstruate in two districts of Malawi, examining both the type of menstrual absorbents used and methods for washing and drying of absorbents. Through a 2019 survey of over 900 households, conducted by trained NGO workers, MHM at the household level is investigated. The study underscores the necessity for enhanced MHM in Malawi, focusing not just on ensuring access to and affordability of suitable menstrual absorbents, but also on advocating for proper washing and drying practices for these absorbents. Specifically, this research paper specifically addresses the following research questions: (1) What are the most common methods of menstrual hygiene management by people who menstruate in Malawi? (2) How are reusable menstrual absorbents washed and dried?

## 3 Methods

### 3.1 Study location

Malawi is a low-income country in south-eastern Africa with a population of 19.9 million (World Bank, 2023a), this is rapidly growing with an annual population growth rate of 2.6% (World Bank, 2023b). 70.1% of the population live below the international poverty line of $2.15/ day (defined in 2017)(World Bank, 2023c), this makes purchasing sanitary products for appropriate MHM a challenge.

Surveys were conducted within 2 districts of Malawi, not disclosed for anonymity purposes, referred to as districts A and B. Districts A and B were selected as part of a wider sanitation and hygiene survey (Hinton et al., *in review*) of communities that had both undergone several Community Led Total Sanitation (CLTS) [CLTS] interventions and subsequently declared as open-defecation free. CLTS is a behaviour change programme focused primarily on ensuring sanitary provision and ending open defecation on a community level, emphasising the community wide and environmental health significance of sanitary provision (Cavill et al., 2015). However, CLTS can also be expanded to address MHM, creating a holistic view of multiple aspects of sanitation and hygiene and emphasising the community wide and environmental health consequences of appropriate MHM (Roose et al., 2015).

### 3.2 Survey methodology

Interviews of a total of 939 households were conducted in 2 districts of Malawi, district A and district B. Interviews were conducted as part of a wider survey on sanitation and hygiene provision (Hinton et al., *In review*). Households surveyed belonged to the service area of 10 selected Healthcare Centres (HCC), six in district A and four in district B. Households within these service areas were selected on a stratified random sampling basis, taking the same proportion of households surveyed within each community (approximately 10-20%). Surveys were conducted by trained staff members in Chichewa and English. The interviews were conducted by trained NGO workers, in collaboration with District and Area Environmental Health Officers, and District Water Development Officers in Chichewa. Data was collected using a mobile based forms which were checked and validated by a verifier. Some of the enumerators used paper forms before transferring the data onto the mobile form later.

Surveys were conducted between the 7th and 25th of July 2019. Trained staff visited households and asked for a member of the household to answer a variety of questions relating to sanitation and hygiene. Regarding MHM, household members were asked “Which type of materials are used by women in this household to collect/absorb menstrual blood?”. Respondents were able to list multiple materials/methods, including the option to say, “don’t know/refuse to answer”. Respondents that seemed too uncomfortable (e.g., being silent for a while, turning around, or diverting the topic to something else) were recoded as “don’t know/refuse to answer”. Those using disposable tissues, e.g., wet wipes, paper towels, or ‘Kleenex’ type of materials, were recorded as “toilet paper”. Respondents that reported using reusable cloths or rags were then asked about management of the menstrual absorbents, including the method for washing and drying from several options. Respondents, therefore, did not necessarily menstruate themselves but, where they could, provided a response regarding the MHM of people who menstruate in the household. Where this was either not relevant or the participant had insufficient information to answer the questions, there was an option for no response to be given. To better understand MHM practices and views within the communities, several focus groups were organised with women groups prior and in parallel to the household surveys. Quotes from respondents have been anonymized.

Informed verbal consent of study participants was obtained prior to their participation. Before proceeding with the questionnaire, the interviewers provided background information on the survey (e.g., objectives, anonymity, length of the interview, etc.) and specified that the respondent could decide to not respond or skip a question or stop the interview at any time. Interviewers introduced themselves, mentioning the name and the organisation for whom they worked as well as where they could be contacted in case the interviewee would have any further questions or concerns after the survey.

### 3.3 Data interpretation

Responses were restricted to within the sample window specified and duplicate responses were removed prior to analysis. The type of MHM was summarised, any response which listed a type of menstrual absorbent was summed to give an estimate for the total number of people who use a given menstrual absorbent product. Results of the survey for district A and B are summarised separately to establish if any major differences between the districts existed.

The number of households without access to appropriate menstrual absorbents was calculated as the sum of households listing toilet paper or leaves as the only menstrual absorbent reported for MHM. The number of households with ‘at risk’ MHM practises was calculated as the sum of the total number of households using reusable menstrual absorbents with inadequate washing practices (households that did not wash menstrual absorbents or washed menstrual absorbents without soap) and the estimated number of households that had appropriate washing practices but inadequate drying practices. The estimated number of households with inappropriate drying was calculated as the total number of households where menstrual absorbents were washed with soap and water multiplied by the percentage of households that dried menstrual absorbents inside.

## 4 Results

### 4.1 Menstrual absorbents used

District A surveyed 733 households whilst 206 households were surveyed in district B, 714 and 187 households provided an answer regarding the menstrual absorbents used in district A and B respectively. The type of menstrual absorbent used by households is summarised in Table 1, multiple answers could be provided. Reusable cloths or rags were listed as a menstrual absorbent used by 79.5% of households in which the participant provided an answer: 81.1% in district A and 73.3% in district B. Disposable menstrual absorbents, such as tampons and pads, were used by 18.6% of households that provided a response: 16.5% in district A and 26.7% in district B. District A also reported that 0.42% and 1.96% of households that reported a method of MHM used toilet paper and leaves respectively, no households reported using these methods in district B.

**Table 1:** Type of menstrual absorbents used by households in district A and B.

| Menstrual product | Number of households in District A using product | Percentage of households in District A using product (%) | Number of households in District B using the product | Percentage of households in District B using product (%) |
| --- | --- | --- | --- | --- |
| Reusable cloth or rags | 579 | 81.1 | 137 | 73.3 |
| Don't know/refuse to answer | 35 | 4.90 | 19 | 10.2 |
| Toilet paper | 3 | 0.42 | 0 | 0 |
| Leaves | 14 | 1.96 | 0 | 0 |
| Disposable (tampons or pads) | 118 | 16.5 | 50 | 26.7 |

In most cases, only one type of menstrual absorbent was listed as being used by households. Of the 716 households that reported using reusable cloths or rags, 699 households (97.6%) reported this as their only menstrual absorbent: 97.4% in district A and 98.5% in district B. 152 of the 168 households (90.5%) that reported using disposable absorbents (tampons or pads) reported these as their only menstrual absorbent, 15 reported using these alongside reusable cloths or rags (8.93%), and 1 household reported using disposable absorbents alongside an unreported method (don’t know/refuse to answer.) A total of 17 (1.81%) households used toilet paper or leaves as menstrual absorbents. Of the 3 households that used toilet paper, 2 reported this as their only menstrual absorbent whilst 1 household reported using toilet paper as well as reusable cloths/rags. Similarly, of the 14 households that reported using leaves, 13 reported that this was the only type of menstrual absorbent used, whilst 1 reported using leaves alongside reusable cloths/ rags.

### 4.2 Washing and drying of menstrual absorbents

The method of cleaning reusable menstrual hygiene products was also evaluated, the results are summarised in Table 2. Overall, 99.0% of households that used reusable cloths or rags in MHM reported that they were cleaned: 99.1% and 98.5% in district A and B respectively. Of those houses cleaning reusable cloths or rags, 90.1% used soap and water (89.0% and 91.1% in district A and B respectively), whilst 9.94% cleaned with water only (10.1% and 8.89% in district A and B respectively).

**Table 2:** Type of washing of reusable menstrual absorbents in district A and B.

| Method of cleaning menstrual absorbent | Number of households in District A | Percentage of households in District A (%) | Number of households in District B | Percentage of households in District B (%) |
| --- | --- | --- | --- | --- |
| Not washed | 5 | 0.9 | 2 | 1.5 |
| Washed with water only | 58 | 10.1 | 12 | 8.89 |
| Washed with soap and water | 511 | 89.0 | 123 | 91.1 |

Overall, 79.7% of households cleaning reusable menstrual products dried the absorbents inside their home and 20.3% dried menstrual absorbents outside, in the open air. 552 households in district A and 128 in district B provided an answer about how reusable menstrual absorbents were dried. 444 (80.4%) and 98 (76.6%) households reported that they were dried inside whilst 108 (19.6%) and 30 (23.4%) households reported that the products were dried outside in the open air in district A and B respectively.

In total, 77 (10.8%) households that used reusable menstrual hygiene products (and reported on the method of cleaning) were inappropriately washing their reusable menstrual hygiene products, either by not washing menstrual absorbents or washing with water only.) 79.7% of households using and washing reusable menstrual absorbents reported drying the products inside their home; an estimated 505 households wash menstrual absorbents with water and soap before drying these inside. Collectively, 582 of the households using reusable cloths or rags report either washing or drying their menstrual absorbents inappropriately; only 18.1% of households using reusable cloths or rags practised appropriate washing and drying.

## 5 Discussion

A major focus in ensuring adequate and equitable sanitation and hygiene is considering the menstrual hygiene management (MHM) of people who menstruate. This is specifically highlighted in SDG6.2, which emphasises the importance of paying special attention to the needs of women, girls and those in vulnerable situations (UN General Assembly, 2015).

Within Malawi, reusable menstrual absorbents are commonly used for MHM; 68.5% of women report using reusable menstrual absorbents whilst 28.5% use non-reusable absorbents and 2.6% have other/no materials (NSO, 2021). Higher levels of reusable menstrual absorbent use are reported in rural areas, by older women, poorer households, and those with a lower level of education (NSO, 2021). To investigate the types of MHM practised in greater details, we evaluated the results of a survey of 939 households conducted across 2 districts in Malawi (Hinton et al., *in review*). These results provide a comparison to the estimations of menstrual absorbent use and provide additional information about the types of absorbent used. Furthermore, crucially, this work also evaluates the cleaning of reusable menstrual absorbents, which can greatly impact their safety.

We investigated MHM by asking what materials are ‘used by women in the household to collect/absorb menstrual blood’. Reusable cloths or rags were the most common method, used by 79.5% of all households that listed a method of menstrual absorbent. This was similar across both the districts investigated: 81.1% of households in district A and 73.3% of households in district B. These findings report a slightly higher level than the 2021 JMP MICS survey (reported 68.5% reusable absorbent use), however, this is likely due to the surveyed communities being in more rural areas (NSO, 2021). These findings are also consistent with further literature, which reports that reusable menstrual cloths are the most common form of menstrual absorbent used in menstrual hygiene management in Malawi (Pillitteri, 2011), similarly to other low-resource countries (Kuhlmann et al., 2017; Tegegne & Sisay, 2014). Where reusable cloths and rags were used in MHM, they were usually the only method of menstrual absorbent used; reusable cloths and rags were the only menstrual absorbent used in 97.6% of households in which they were used.

Reusable menstrual absorbents made of cloths and rags are less absorbent than commercially available sanitary pads (Foster & Montgomery, 2021), this can prove ineffective in managing menstrual bleeding, sometimes resulting in blood stains on clothes that result in embarrassment, stigmatisation, anxiety, and shame (Kambala et al., 2020; McMahon et al., 2011). Poor absorbency and staining of clothes have been reported as a particular concern for those attending school, with accounts of people who menstruate dropping out of school due bullying around these issues (Tegegne & Sisay, 2014). The fear of ridicule has also been reported to decrease the confidence of people who menstruate in school settings (Vaughn, 2013). In addition, cultural considerations surrounding menstrual blood are an important factor in appropriate MHM. There is a particular concern among by people who menstruate in Malawi about menstrual blood not being seen for fear of menstrual blood being used in witchcraft (Pillitteri, 2011). Within Malawi, 12.7% ‘of women age 15-49 years reporting menstruating in the last 12 months did not participate in social activities, school or work due to their last menstruation’ (NSO, 2021). This further emphasises challenges resulting from menstrual absorbents with inadequate absorbency. In cases where people who menstruate have access to more discrete menstrual products, which cannot be seen under clothing, some cultural beliefs and practises associated with menstruation, such as being unable to be in the same environment as a boy or man, appear to be dying out (Kambala et al., 2020). There are some reusable menstrual products, made from cloth, that offer improved absorbency and protection to using strips of cloth and rags, these are either locally made or commercially available reusable pads (House et al., 2012.). This survey did not specify the type of reusable cloths or rags used as menstrual absorbents; it may be possible that some users utilise purpose-made reusable pads which may provide better absorbency than strips of cloths or rags. Further studies could investigate the different type of reusable cloth menstrual absorbents used in Malawi and how improved reusable menstrual absorbents can be promoted.

Disposable menstrual absorbents were the second most common method, used by 18.6% of all households (16.5% in district A and 26.7% in district B). Disposable menstrual absorbents have greater absorbency than cloths and rags (Foster & Montgomery, 2021) and are often a preferred method of MHM (Hennegan et al., 2016) with reports of young girls in particular preferring disposable menstrual pads (Kambala et al., 2020). However, affordability poses a major limitation to the wider use of disposable menstrual absorbents (Boosey et al., 2014; Kambala et al., 2020; Vaughn, 2013) and concerns have also been raised about enabling safe management of used disposable menstrual products (House et al., 2012.; Kambala et al., 2020). Furthermore, most adults who menstruate report preferring reusable cloths or rags as these are a traditional form of MHM (House et al., 2012; Kambala et al., 2020). Malawi implemented a tax exemption (formally, a 16.5% levy) on menstrual pads from the 1^st^ of April 2022 (The Star, 2023), whilst this may help to improve access to disposable menstrual pads, the cost of sanitary pads is still high. In most cases where disposable menstrual absorbents were used, these were the only type of menstrual absorbent (90.5%), however, 8.93% of households using disposable menstrual absorbents reported using them alongside reusable cloths and rags. This may be due to the high cost of disposable menstrual absorbents, with reusable cloths and rags being used alongside disposable methods despite a preference for disposable products (Kambala et al., 2020).

Financial barriers are not only due to the affordability of products, access to financial resources is also a barrier to appropriate MHM with men often not providing access to money to purchase menstrual hygiene products (McMahon et al., 2011; Shah et al., 2023). This was echoed by participants who emphasised that there is a taboo of asking male household members for money to buy menstrual hygiene products.

Appropriate disposal of menstrual absorbents in also a concern, particularly in the case of disposable menstrual absorbent use. There is a wide lack of appropriate disposal of menstrual waste worldwide with menstrual absorbents typically disposed of by burying, burning or depositing absorbents in garbage or toilets (Kaur et al., 2018). It is recommended that discarded sanitary products should be incinerated for appropriate disposal (Parthasarathy et al., 2022). Appropriate disposal of menstrual absorbents is important for ensuring environmental and community level health due to inappropriate disposal resulting in contact with disease causing pathogens from menstrual products (Kaur et al., 2018; Parthasarathy et al., 2022). Within Malawi, household waste is commonly disposed of within pit-latrines (Hinton et al., 2023), presenting a potential concern for groundwater contamination from disposed menstrual absorbents.

District A also reported that a small number of households used toilet paper and leaves as menstrual absorbents (0.43 and 2.00% of households respectively). These methods were not reported in district B, likely due to the smaller sample size of surveyed households within district B. Leaves have been reported as a menstrual absorbent (House et al., 2012; Vaughn, 2013) but have unreliable levels of absorbency (Foster & Montgomery, 2021) and present a high risk of contamination as well as being difficult and uncomfortable to use (House et al., 2012). In total, 15 households (1.60%) reported using only leaves or toilet paper for MHM, these were classed as households with inadequate access to menstrual absorbents for the purpose of this study. Most households had access to, at least basic, menstrual absorbents.

With people who menstruate mostly using reusable cloths and rags for MHM, washing menstrual absorbents is critical in ensuring appropriate menstrual hygiene in Malawi. Das et al. (2015) (Das et al., 2015) found that women using reusable menstrual absorbents were twice as likely to report a urogenital infection than women using disposable menstrual absorbents, whilst this may have been influenced by other hygiene practises or socioeconomic factors, it highlights the impact of menstrual absorbents on reproductive health. The washing and drying of reusable menstrual absorbents have been identified as a critical consideration (Mahajan, 2019), adequate washing and drying is central to the prevention of microbial growth on the reusable absorbent which may otherwise result in infections (Das et al., 2015; Torondel et al., 2018). Washing reusable menstrual products with water and but not soap has been shown to be associated with more symptomatic urogenital infections (Das et al., 2015). For the purposes of this study, reusable menstrual absorbents must be washed with soap to be classed as adequate MHM, this is consistent with other literature (Hennegan et al., 2016; Ramaiya & Sood, 2020). 99.0% of households that used reusable cloths and rags cleaned the menstrual absorbent, 90.1% of those households used water and soap to clean the cloths whilst 9.94% used only water. Whilst most households reported the use of soap in cleaning menstrual absorbents, it is difficult to measure the actual levels of soap usage in washing as the survey relied on self-reporting. Social perception is an important consideration in self-reported behaviours and may result in an overestimation of such practises (Hennegan et al., 2016).

Drying is also an important practice in the management of menstrual absorbents. Drying in the sun is a method of minimising microbial growth on menstrual absorbents due to the microbiocidal effects of UV light (Bloomfield et al., 2011; Das et al., 2015; House et al., 2012.; Mahajan, 2019; Torondel et al., 2018). Overall, 79.7% of households using, and washing, reusable menstrual products dried the absorbents inside and 20.3% dried menstrual absorbents outside. Despite the benefits of drying menstrual absorbents under the sun, most households dried menstrual absorbents inside the house, this may be due to the stigma surrounding menstrual hygiene as drying menstrual absorbents in the sun may cause embarrassment due absorbents being more easily seen (Averbach et al., 2009; Kuhlmann et al., 2017). This is similar to MHM techniques in other countries within Sub-Saharan Africa; within Uganda, a survey of schoolgirls in rural Uganda found that 41.6% of users dried absorbents outside with 47.4% of users drying menstrual absorbents hidden inside (such as under beds) where drying is especially limited, raising concerns for infection risk (Hennegan et al., 2016). Cultural attitudes are essential in consideration of washing and drying practices, as one of the focus group participants shared: “is a taboo to dry MHM materials (clothes/rags) outside the house, they should be dried inside the house” (personal communication, 18/03/2019). This is particularly relevant considering cultural attitudes and stigma associated with menstruation in Malawi (Pillitteri, 2011). Our results suggest that the biggest barrier to the appropriate management of reusable menstrual absorbents was not access to materials (such as soap) but rather inappropriate drying, we suggest that this may be linked to the stigma surrounding MHM.

Overall, we estimate that 81.9% of households using reusable menstrual absorbents had inappropriate washing or drying methods, including households not washing menstrual absorbents, washing with only water, and/or drying menstrual absorbents inside. Nonetheless, some women in the community know about health risks associated to improper washing and drying of reusable materials. One of the focus group participants shared that “rags/clothes which women use as vaginal pads should be washed with soap and air dried outside the house so as to get rid odour and infection (sic)” (personal communication, 18/03/2019).

Whilst this study provided a large-scale evaluation of MHM across two districts in Malawi, and an indication of MHM practises and challenges, the study also has key limitations. Surveys were conducted by household members as part of a wider survey on sanitation, therefore, in some cases, a household member who did not menstruate provided a response on the method of MHM used by menstruating household members. To minimise this leading to false responses, there was an option for interviewees to provide no response or state that they did not know. Future work would benefit from specifying whether the interviewee menstruated themselves.

Spatial variation may be another critical consideration in evaluating MHM across Malawi. Whilst this survey found similar results in both district A and B, it only surveyed communities within 2 districts making it hard to draw national conclusions. There may be significant spatial variation between districts that is not represented in these results. Notably, there may be differences between the most common MHM in urban and rural contexts, with people living in urban areas often having greater access to sanitation (Rossouw & Ross, 2021), future work could provide a more comprehensive overview of MHM across Malawi, accounting for variation between rural and urban communities. This was mentioned by focus groups who highlighted geographical challenges to accessing commercial menstrual hygiene products with users closer to trade centres speaking of using single use products more frequently. Furthermore, the two districts identified for the survey presented here were evaluated due to previously being declared open defecation (Hinton et al., *in review*), it may be that the districts presented here are not representative of MHM across Malawi due to previous investment into gender-sensitive sanitation and hygiene programmes in the district to achieve SDG6.2.

Our study indicates that, despite improvements in MHM in Malawi and government measures to promote safe MHM, a large proportion of people who menstruate have inadequate access to menstrual absorbents and do not practise safe cleaning of these products. To further understanding of MHM in Malawi, future country-wide surveys including Censuses, DHS, and MIS surveys, conducted by the Government of Malawi and in collaboration with partner organisations, should include space for more information on MHM.

## 6 Conclusion

Through evaluating menstrual hygiene management (MHM) across two districts in Malawi and surveying over 900 households, we find that reusable cloths or rags are the most used menstrual absorbent product, used by 79.5% of households. 1.6% of households surveyed did not have access to any appropriate menstrual absorbents and instead reported using only toilet tissue or leaves.

Investigating the nature of washing and drying of reusable menstrual absorbents provided an indication of hygiene challenges of reusable products. Whilst, in most cases, these are appropriately cleaned with soap and water (90.1% of households), most households did not dry the menstrual absorbents outside in sunlight (only 20.3% of households dried menstrual absorbents outside) which can be important in preventing microbial growth. Overall, we estimate that, despite most households having access to at least a basic menstrual absorbent (reusable cloths/rags or a disposable absorbent), appropriate management is holding back adequate MHM. Approximately 81.9% of households did not implement appropriate washing or drying of menstrual absorbents. We suggest that the stigma associated with menstrual hygiene may be preventing appropriate MHM with important public health consequences, particularly around the prevention of urogenital infections. Education and behaviour change communications campaigns around appropriate usage should highlight the importance of appropriate MHM.

Further studies into MHM on a national level in Malawi will be essential to better understand the status of menstrual hygiene in Malawi; enabling Malawi to reach SDG6.2 access to adequate and equitable hygiene for all. Additionally, further studies should explore barriers to appropriate MHM practice within Malawi, investigating social, cultural, and economic challenges.

## Data Availability

Confidential data were provided by the Government of Malawi and CARE. All data summarised is provided here.

## Acknowledgements

We would like to acknowledge the work of CARE in enabling this research.

## Consent

Informed consent was obtained from all subjects involved in the study. All data collected was in line with the Government of Malawi ethics and was agreed with each participant.

## Data availability

Confidential data were provided by the Government of Malawi and CARE. All data summarised is provided here. Additional data was obtained from publicly surveys including US Aid Demographic Health Surveys (DHS) Government of Malawi Census Data and US Aid Malaria Indicator Surveys (MIS).

